# NeuroDev: etiology and experience of neurodevelopmental disorders in Kenya and South Africa

**DOI:** 10.64898/2026.04.30.26351947

**Authors:** Patricia Kipkemoi, Emily O’Heir, Mutaz Amin, Sarah L Stenton, William Baddoo, Harrison Brand, Zandre Bruwer, Sam Bryant, Eunice Chepkemoi, Björn Christ, Emma Eastman, Claire Fourie, Jack M. Fu, Alice Galvin, Stacey Hall, Heesu Ally Kim, Fatima Khan, Collins Kipkoech, Martha Kombe, Rachael Mapenzi, Brigitte Melly, Celia van der Merwe, Beatrice Mkubwa, Serini Murugasen, Katini Mwangasha, Paul Mwangi, Samuel Mwasambu, Alfred Ngombo, Javan Nyale, Ikeoluwa Osei-Owusu, Jessica E. Ringshaw, Kathryn A Russell, Kaitlin E. Samocha, Alba Sanchis-Juan, Moriel Singer-Berk, Grace E. VanNoy, Michal Zieff, Michael E. Talkowski, Anne O’Donnell-Luria, Christina Austin-Tse, Charles R. Newton, Amina Abubakar, Kirsten A. Donald, Elise B. Robinson

**Affiliations:** Neuroscience Unit, KEMRI-Wellcome Trust, Center for Geographic Medicine Research Coast, Kilifi, Kenya; Institute of Human Development, Aga Khan University, Nairobi, Kenya; Center for Genomic Medicine, Massachusetts General Hospital, Boston MA, USA; The Broad Institute of MIT and Harvard, Cambridge MA, USA; Department of Neurology, Harvard Medical School, Boston MA, USA; Department of Paediatrics and Child Health, Red Cross War Memorial Children’s Hospital, University of Cape Town, Rondebosch, South Africa; Department of Psychiatry, University of Oxford, London, UK; Division of Genetics and Genomics, Boston Children’s Hospital, Boston MA, USA; Neuroscience Institute, University of Cape Town, Groote Schuur Hospital, Observatory, South Africa

**Author notes:** Correspondence to Drs. Abubakar, Donald, and Robinson. Joint first authors (Patricia Kipkemoi, Emily O’Heir).

## Abstract

The NeuroDev study, conducted in Kenya and South Africa, is a large-scale clinical, genetic, and epidemiologic characterization of neurodevelopmental disorders (NDDs) on the African continent. NeuroDev assessments capture birth, demographic, and developmental history; cognitive and behavioral outcomes; and physical health variables. DNA samples are collected for exome sequencing and clinical genetic analysis. This paper presents novel data from 521 children with NDDs, 739 of those children’s parents, and 255 unrelated, typically-developing children. The analyses offer unique genetic and phenotypic characterizations of NDDs in two African countries and underscore the importance of including underrepresented populations in NDD research. Ultimately, 107 children with NDDs from the NeuroDev cohort (22.1%) had likely pathogenic or pathogenic variants in established NDD genes. High rates of genetic diagnosis were associated with high rates of environmental risk factors for NDDs. All data, materials, and measures generated from this study are publicly available through the US National Institute of Mental Health.

## Main

Following decades of efforts to improve child health outcomes, far more children in African settings are living past their fifth birthdays and are being diagnosed with neurodevelopmental disorders (NDDs).^1,2^ While they number in the millions, these children are rarely included in NDD research cohorts. The experience and etiology of NDDs in the African context is unstudied at scale.

The NeuroDev Project was built to address this gap, and to characterize the clinical, genetic, and environmental architecture of NDDs in Kilifi, Kenya and Cape Town, South Africa.^3,4^ NeuroDev primarily enroll children who meet diagnostic criteria for global developmental delay/intellectual disability (GDD/ID) and/or autism spectrum disorders (autism). We enroll as many of those children’s parents as are available and willing to participate, as well as unrelated, age- and ancestrally-matched children who do not have developmental disorders. Here we present analyses of data from 1515 study participants, 521 of them children with NDDs.

Children enrolled in NeuroDev complete a four hour neuro-medical assessment capturing birth, demographic, and developmental history; cognitive and behavioral outcomes; and physical health variables. The NeuroDev assessments are designed for use in lower and middle-income countries (LMICs) and are freely available for use. In Kilifi, NeuroDev families completed the assessments in Kiswahili or Kigiriama, in Cape Town, English, isiXhosa, or Afrikaans. All participants provided DNA samples extracted from blood, we also piloted saliva collection of a subset of participants, and 398 South African samples were processed for generation of cryopreserved cell lines (CPLs). Many South African families also consented to share photos of their children. These photos address another significant gap in representation - almost all existing medical reference photographs for NDDs come from North America or European communities.^5^

Analyses presented here capture the clinical picture of NDDs in Kilifi and Cape Town, genetic diagnoses made in the context of African genetic diversity, and location-specific environmental risk factors for NDDs. All NeuroDev data can be accessed through the National Institute of Mental Health (NIMH).

## Results

### The NeuroDev cohort

A total of 1515 participants comprise the NeuroDev Wave 1 cohort: 521 children with NDDs, 457 mothers, 282 fathers and 255 unrelated children without NDDs. Child controls were matched to child cases on self-reported ancestry, geographic location and age. Children enrolled in the study were between the ages of 2 and 17 years. Those with NDDs were disproportionately male (71.4%); the typically developing children were not (48.6% male; Table 1). The mean age of children with NDDs enrolled in Cape Town was 5.6 years, substantially younger than those enrolled in Kilifi (11.7 years). This difference reflects site-specific recruitment strategies. In Cape Town, children and families were recruited from the developmental medicine clinics at Red Cross Children’s Hospital and Tygerberg Hospital. In Kilifi, participants were recruited from clinics, special needs schools, and some had been recontacted from previous studies of ID and epilepsy in the region.

**Table 1.**
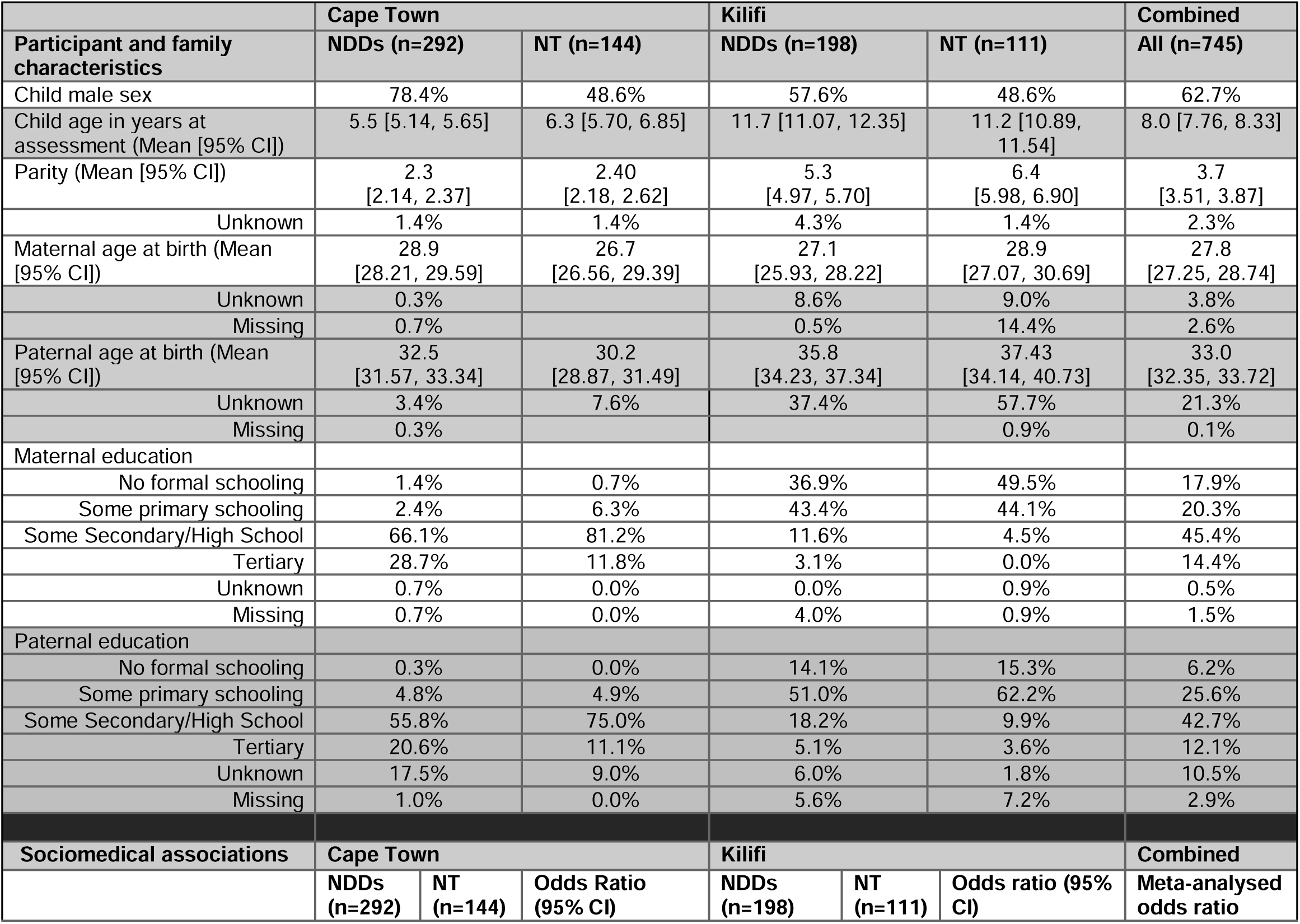
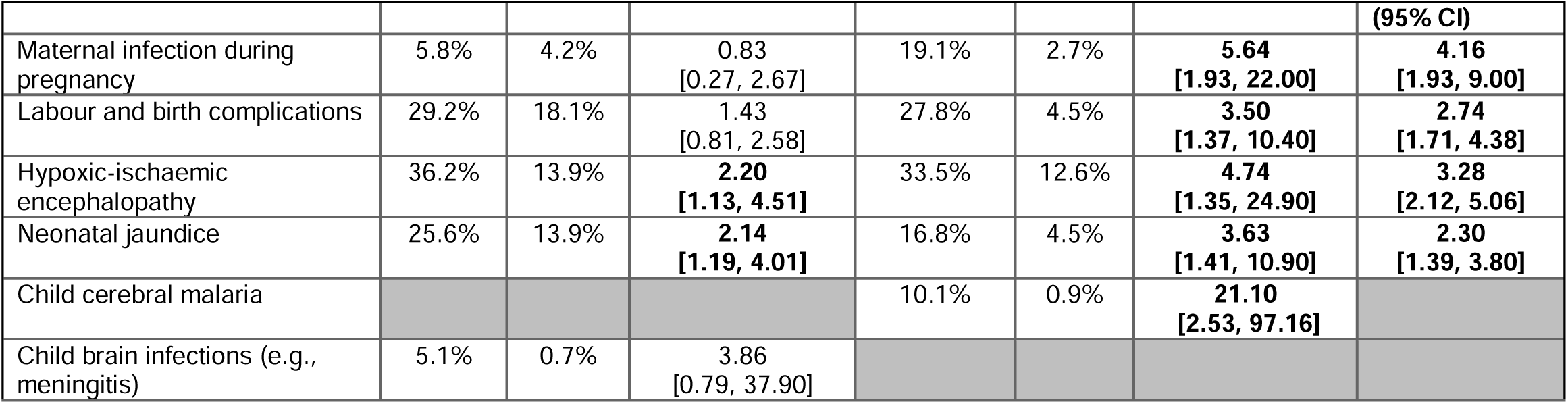
- The NeuroDev Cohort, non-genetic factors associated with NDDs

There are many regional differences between Cape Town and Kilifi (**Table 1**). Kilifi is a rural county, and most adults complete their formal schooling in primary school. Most people living there have parents and grandparents who were born in Kenya, and who speak the local languages. Cape Town is an urban area with high rates of adult literacy, and expansive diversity in educational, medical and socioeconomic opportunity. It is an international city in terms of cultural and ancestral diversity. Most NeuroDev parents from Cape Town completed high school, and approximately one quarter of NDD parents completed college. This is an overrepresentation of college-level education in Cape Town and may point to genetic literacy as a driving motivation to enroll in NeuroDev South Africa. NeuroDev South Africa offered genetic testing with return of clinical findings.^6^ NeuroDev Kenya did not offer feedback of NDD genetic diagnoses. This decision reflected wishes of the Kilifi community, expressed during structured conversations with ethicists on the NeuroDev Kenya team. These and other ethical conversations continue with Kilifi community members and NDD stakeholders in a community-based participatory research framework.

All data collection in NeuroDev is conducted by team members fluent in the participants’ preferred languages. Trained research nurses and research assistants were responsible for the recruiting, consenting, assenting and assessment of participants in Cape Town. Trained field team members were responsible for recruiting, consenting and assenting participants in Kilifi, where assessments were then conducted by research nurses or clinical officers. We used the University of California, San Diego Brief Assessment of Capacity to Consent (UBACC) to measure parents’ understanding of the study prior to consent.^7,8^ Specifically trained research staff used the UBACC to identify parents who may require a more comprehensive evaluation of decisional capacity and enhanced consent procedures.

Key characteristics of the NeuroDev cohort (NDD and neurotypical (NT)) are presented in **Table 1**. As described above, the cohort demographics reflect many common regional features. For example, average family size is larger in Kilifi than in Cape Town. In Kilifi, it is common for adults to estimate their age within a window, rather than mapped to a specific birthday. In the table above, this circumstance is reflected as “unknown” age, where non-response is coded as “missing.”

**Table 1** also presents non-genetic factors associated with NDDs in NeuroDev. We estimated unadjusted and adjusted logistic regression models of the association between non-genetic factors and NDDs. In Cape Town, children with NDDs were more likely to have experienced hypoxic-ischemic encephalopathy (HIE) (OR 2.20 [95%CI 1.13, 4.51]) and neonatal jaundice (OR 2.14 [95%CI 1.19, 4.01]) than NT children. In Kilifi, each of the following was significantly associated with NDDs: maternal infection during pregnancy (OR 5.64 [95%CI (1.93, 22.00]); labor and birth complications (OR 3.50 [95%CI 1.37, 10.40]; HIE (OR 4.74 [95%CI 1.35, 24.90]); neonatal jaundice (OR 3.63 [95%CI 1.41, 10.90]; and childhood cerebral malaria (OR 21.10 [95%CI 2.53, 97.16].

NDD families had fewer total children in both Cape Town (OR 0.77 [95%CI 0.62, 0.97] and Kilifi (OR 0.87 [95%CI 0.78, 0.97]) compared to NT families, consistent with global observations of “stoppage” following the birth of a developmentally disabled child.^9^ In Kilifi, children with NDDs were more likely to be delivered in a hospital than at home (OR 0.27 [95%CI 0.13, 0.53]). This may reflect experience or identification of challenges during the pregnancy.

**Figure 1**. Human Phenotype Ontology (HPO) terms reported in ≥5 NeuroDev probands. Individual proband counts are displayed on the bars, proportion of the respective cohort is displayed on the x-axis. *Medical records were only available for the South Africa cohort. HPO terms extracted from medical records are not available for the Kenyan cohort (denoted as N/A).

**Figure 1.**
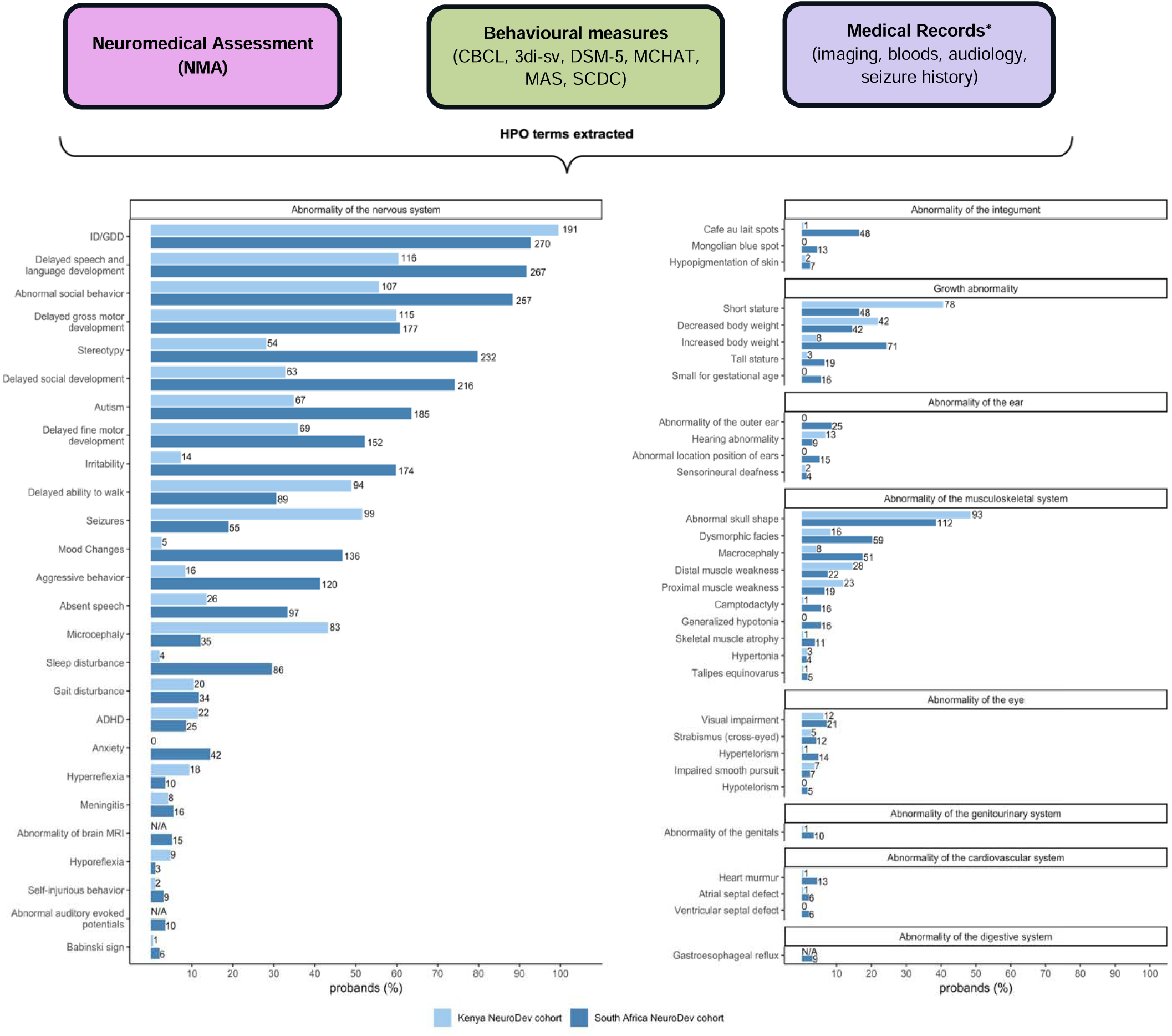
Clinical Spectrum of NDDs in NeuroDev

### Clinical characteristics of the NeuroDev case cohort

The NeuroDev phenotype battery includes demographic assessments, a site-specific asset index detailing socio-economic status, neuromedical assessments capturing birth history, cognitive assessments and behavioral assessments (**Figure 1**). The cognitive assessments included the Raven’s Progressive Matrices^10^ (RPM; standard for ≥12 years of age, colored for 6-11 years of age), which measure nonverbal reasoning and the Molteno Adapted Scales^11^ (MAS) to evaluate developmental delay. The RPM has acceptable internal consistency in both of our study settings, with Cronbach’s alpha ranging from 0.70 and 0.84. ^12,13^ The MAS was developed in South Africa and shows moderate to high correlation with the Bayley Scales of Infant and Toddler Development and good diagnostic accuracy for global developmental delay.^14,15^ We adapted the MAS for use in Kilifi. For example, we removed a question about riding a tricycle, as it was not culturally appropriate, and replaced it with a question about standing on tiptoes.^4^ At both sites, all participants 6 years and older attempted the age-appropriate RPM, and those who could not complete the standard RPM attempted the colored version of the RPM. Those under age 6 years, and those who could not complete an age-appropriate or colored RPM, were assessed using the MAS. All NeuroDev phenotypes were annotated with Human Phenotype Ontology (HPO) terms, a structured ontology which aids in integrating clinical data and conducting clinical genetic analysis.

Primary behavioral diagnoses in NeuroDev (e.g. autism; attention deficit hyperactivity disorder (ADHD)) were based on DSM-5 criteria and trained clinician judgement. We supplement those diagnoses across all children using several behavioral measures: the Child Behavior Checklist^16^ for ages 1½-5 and 6-18 years (CBCL1/½-5 and CBCL 6-18); the Swanson, Nolan, and Pelham Rating Scale^17^ (SNAP-IV) to measure ADHD symptomatology; the short version of the Developmental, Dimensional and Diagnostic Interview^18,19^ (3di-sv) and the Social Communication Disorders Checklist^20^ (SCDC) to measure characteristics of autism and other behavioral domains. As described in Kipkemoi et al. we modified several of those measures to better match NeuroDev families’ spoken languages and lived environments.^4^

The distribution of human phenotype ontology (HPO) terms among NeuroDev children with NDDs, and the measures from which those terms were derived, are presented in Figure 1. Almost all probands (95%) met criteria for ID or GDD, and approximately half (52%) met criteria for autism. A small number met criteria for autism alone (n=13, 2.7%). An additional small number did not meet criteria for ID/GDD or autism, and were diagnosed with specific learning disabilities, communication disorders, and/or ADHD (1.9%, n=9), additional diagnoses meeting criteria for NeuroDev case inclusion. Age-appropriate RPMs could not be completed by 44.5% of NeuroDev children with NDDs. Children <6 years of age, and those unable to complete the age-appropriate RPM were offered the MAS. An additional 26.0% of individuals with NDDs were unable to fully complete the MAS. NeuroDev children meeting criteria for both autism and ID/GDD were least likely to fully complete the MAS.

In total, 58 HPO terms, spanning multiple organ systems, were extracted across clinical assessments and reported in ≥5 probands (median 13 terms per proband, range 1-45; **Figure 1**). Autism, and autism-associated traits (e.g. repetitive mannerisms), were more frequent in the South African cohort. Children with autism were more likely to be ascertained in NeuroDev South Africa than NeuroDev Kenya, a reflection of the patient population at the recruitment sites. Autistic children seen in NeuroDev-affiliated clinics in Cape Town typically meet criteria for ID and often have high behavioral support needs. Primary ID with syndromic features was more likely to be ascertained in Kenya, which is reflected in greater rates of microcephaly and seizures in Kenyan case individuals. As discussed below, the Kenyan cohort also features a higher proportion of genetic diagnoses.

Many differences between the Kenyan and South African cohorts reflect study- and/or context-specific factors. For example, hyperpigmentation (e.g. cafe au lait spots and congenital dermal melanocytosis) was more frequently reported in the South African cohort. This at least partially reflects the younger average age of the South African children as congenital dermal melanocytosis typically resolves by early childhood. Small for gestational age (SGA) was also reported more often in NeuroDev South Africa than in NeuroDev Kenya. This difference likely reflects variation in documentation. In South Africa, birth and early growth is routinely documented in hand-held “road to health” booklets. By comparison, fetal monitoring in the antenatal period is difficult in Kilifi, and it is difficult to reliably assign gestational age at birth, impeding diagnosis of SGA.

### Ancestry and rare variant counts

Whole exome sequencing and genome-wide common variant data were generated for each individual in NeuroDev. We analyzed the genotypes of 472 unrelated NeuroDev children with NDDs alongside genotypes of 1307 individuals from the African Genome Variation Project (AGVP; **Figure 2**).^21^ In principal components analysis, NeuroDev participants of predominantly African descent tended to cluster with geographically proximate African populations. For example, NeuroDev South African individuals with lower levels of admixture clustered primarily with southern African populations, including the Zulu and Sotho, while NeuroDev Kenya samples clustered closely with eastern African populations such as the Baganda and Barundi. As expected, there was greater admixture and ancestral heterogeneity, in the NeuroDev South African cohort.

**Figure 2.**
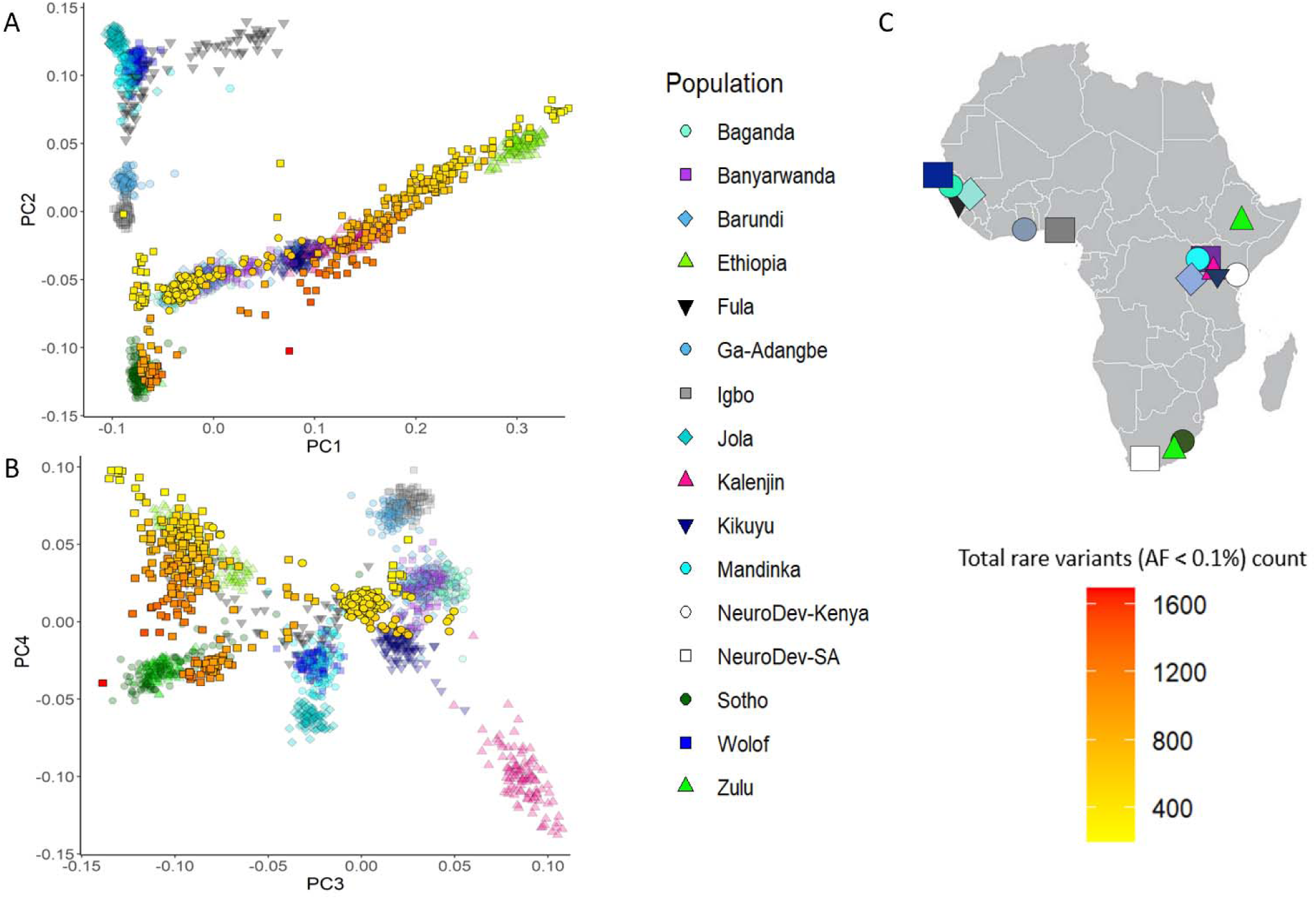
Rare variant counts by genetic ancestry in NeuroDev. **A.** African sub-continental PCA plots of NeuroDev Kenya (circles) and South Africa (squares) case individuals projected onto the first 4 PCs of the AGVP panel. **B.** Rare variant count in NeuroDev case individuals is indicated by yellow to red color gradient within the shapes. **C.** Map of geographic populations represented in AGVP, a within-Africa reference panel.

Combining NeuroDev genotyping and sequencing data, we were able to map genetic ancestry to rates of rare (allele frequency <0.1% in gnomAD v4.1) genetic variation in NeuroDev participants. This variation is overwhelmingly benign, reflective of variably increased genetic diversity among African populations. Rare variant counts were indeed correlated with genetic ancestry in NeuroDev, as well as with maternal grandmother spoken language (**Supplemental Figure 1**), which aligns with genetic ancestry in the NeuroDev cohort.^4^ Rare variant rates displayed a clear gradient within the South African cohort. Notably, individuals from South Africa whose genetic ancestry clusters with Ethiopian populations had lower rare variant counts compared to those clustering with indigenous South African groups like the Sotho and Zulu. This reflects lower out-of-Africa admixture among the Sotho and Zulu.

### Rare variant burden analysis across different ancestries

Compared to other neurodevelopmental and rare disease cohorts sequenced by the Broad Center for Mendelian Genomics (CMG), NeuroDev participants carried the highest numbers of rare loss-of-function (LoF), missense/inframe indel, and synonymous variants (**Figure 3a**). This is a combined function of a) Africans having the greatest genetic diversity of any global population and b) limited representation of individuals of Eastern- and Southern- African ancestry in reference databases like gnomAD.^22^ A larger number of rare variants carried by an individual results in more variants returned by algorithmic clinical genetic analysis programs, like *seqr*, and increases the number of variants requiring manual clinical review.^23^ In comparing the *seqr* filtration outputs (returned variant counts) for probands of different genetic ancestries, we indeed observed higher numbers of rare variants returned in South African and Kenyan NeuroDev probands compared to rare disease probands from other genetic ancestry groups, especially when one or both parents were unavailable for variant phasing (**Figure 3b**). Parent-child trio data renders variant return more consistent across ancestries, though trios are often disproportionately challenging to enroll in LMIC contexts. For example, in Kilifi, many fathers work in larger towns or cities away from the family home. This is often especially true for parents of children with NDDs, as there are increased costs of caring for their child.^24^

**Figure 3.**
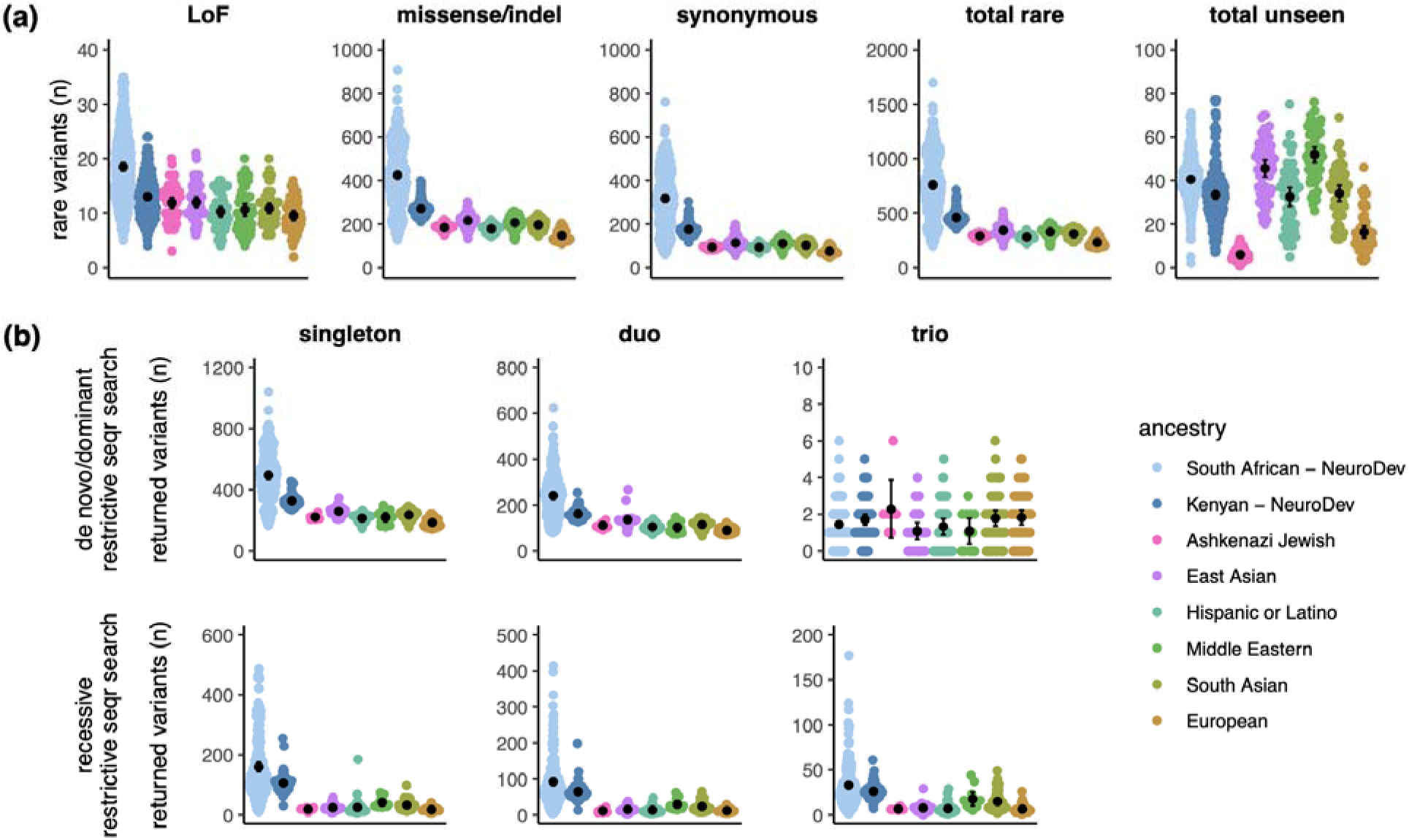
Rare variant counts by genetic ancestry. **a)** Number of rare variants (allele frequency <0.1% in gnomAD v4.1) per proband across protein-coding genes by ancestry and predicted consequence. Total = LoF + missense/inframe indel + synonymous. Unseen = unseen in gnomAD v4.1. Samples included in analysis = 291 South African, 192 Kenyan, 50 Ashkenazi Jewish, 50 East Asian, 50 Hispanic or Latino, 50 Middle Eastern, 50 South Asian, 50 European. **b)** Number of variants returned by *seqr* in *de novo*/dominant and recessive searches per proband by family structure and genetic ancestry. Samples included in analysis = 179 South African, 64 Kenyan, 7 Ashkenazi Jewish, 25 East Asian, 43 Hispanic or Latino, 12 Middle Eastern, 50 South Asian, 50 European.

### Exome sequencing analysis results

Fifty-six (19.2%) South African cases and 51 (26.6%) Kenyan cases were solved with pathogenic (P) or likely pathogenic (LP) variants impacting established NDD genes, contributing to an overall diagnostic yield of 22.1% (**Figure 4a**). The diagnostic variants included LOF (nonsense, frameshift, splicing), missense, inframe indels, and CNVs (**Figure 4b**). The majority (86%) of these variants were either *de novo* or presumed *de novo* when complete parental sequencing data was not available. Representative diagnostic variants and photographs of probands with known syndromic NDDs will be shown in the published manuscript, and variant details for the full cohort are included in Supplementary Table 1. In South Africa, informed consent for the return of study findings was obtained, and P and LP variants were disclosed to participants during in-person genetic counselling sessions, following clinical confirmation of the research findings. For participants with negative findings, including those with no candidate variants or only VUS, negative results were communicated via telephonic consultation, and written summaries were subsequently provided to both the family and the managing clinician.

**Figure 4.**
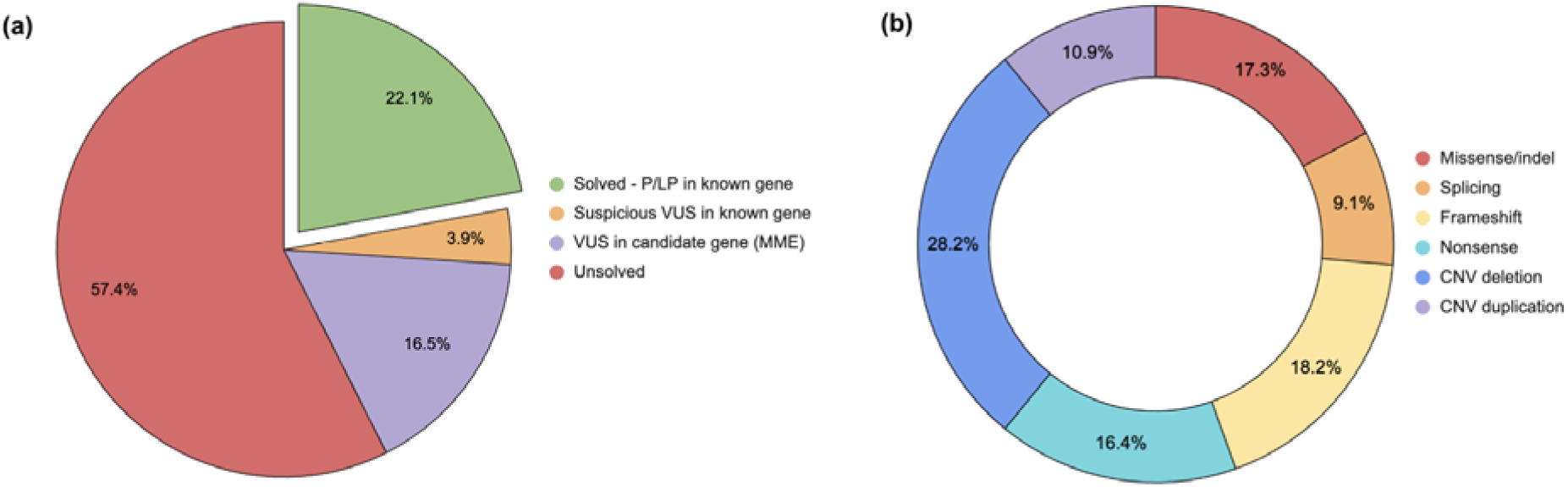
Exome sequencing analysis results summary. **a)** Overall diagnostic rate of the NeuroDev cohort**. b)** Breakdown of diagnostic variants (n = 110) by variant type.

An additional 19 probands (3.9%) had highly suspicious VUS in established NDD genes. These variants were considered to be likely disease-causing, but currently lack sufficient evidence to be classified as P/LP using the ACMG/AMP guidelines. These VUS might be reclassified in the future, if additional functional and/or case-level evidence emerges, potentially leading to more returnable findings.

In addition to identifying causal and likely causal variants in known NDD genes for several African families, NeuroDev participants have contributed to the discovery of more than 30 novel NDD genes. A total of 80 cases had VUS in genes that were not associated with a well-established NDD at the time of analysis, and all candidate genes were submitted to the Matchmaker Exchange platform (MME).^25^ Matches were made for 32 of the submitted cases, each of which will be included in collaborative case series reports describing these emerging gene-disease associations.

NeuroDev collected clinical photographs from case children to improve recognition and characterization of genetic syndromes in African populations. For several NDDs, these images represent the first photographed cases in African-ancestry individuals. Images of 212 children with NDDs and 90 neurotypical children were shared with Face2Gene, an AI tool that can identify genetic syndromes through photographs and is free to use for clinicians worldwide.^26^ NeuroDev photographs aim to improve the performance of its algorithm for African-ancestry populations.

## Discussion

NeuroDev highlights the extent to which clinical care and research into NDDs must be context specific. Clinical characterizations in NeuroDev were tailored to language, culture, and resource availability; genetic analysis strategy shifted in response to continued under-representation of African-ancestry populations in genomic research; sociomedical findings suggested substantial differences in the composition and interpretation of non-genetic risk factors for NDDs in international settings.

NeuroDev probands carry more rare genetic variants than individuals with NDDs from other ancestral groups, reflecting both increased genetic diversity and the underrepresentation of African ancestry in large genetic reference databases such as gnomAD.^27^ While several recent efforts, including the release of gnomAD v.4.1.0, include more diverse populations, the disparity remains and is particularly relevant to people of Eastern- and Southern-African ancestry.^28^ Genetic analyses rely heavily on correctly estimated allele frequency, and the representation gap has immediate clinical implications for genomic medicine. Clinical genetic analyses in NeuroDev required more time and manual review, particularly in the absence of complete parent-child trio data, which slows the diagnostic process. Inclusion of global populations in genomic research continues to be necessary to reduce health disparities.

In South Africa, returning individual genomic results from NeuroDev provided families with diagnoses that would otherwise have been inaccessible, and often offered information about clinical prognosis and potential therapeutic interventions. Families valued counselling with regard to recurrence risk, and it influenced reproductive decision-making. Return of results continues at NeuroDev’s site in South Africa, and researchers in Kilifi are exploring mechanisms of returning results to interested families.

There is also likely to be setting-specificity in the landscape of non-genetic risk factors for NDDs. Reduced healthcare access and resources in LMICs contribute to a higher incidence of birth complications and hypoxic insults, and pre- and peri-natal infections differ in their average composition around the globe.^29^ We observe a strong association to prenatal infection in the NeuroDev Kilifi cohort. This and other environmental association to NDDs should be examined in detail to better understand which types of infections - perhaps those more common in equatorial environments - may create more risk for NDDs.

NeuroDev data capture the experience and etiology of NDDs in two settings under-represented in the research of neurodevelopmental genetics. The settings necessitated bespoke approaches to data collection and characterization, resulting in a unique and high-quality data resource for the global community. Methodological limitations in NeuroDev include dependence on retrospective recall. Recall bias in mothers of children with NDDs, especially those with high support needs, may be present, and these mothers may recall past exposures differently than mothers of neurotypical children.^30,31^ The socio-medical associations described are likely to include influence from residual confounding and, as discussed above, should not be assumed to have a causal relationship to NDDs.

Major advances have been made in the study of NDDs, with studies of NDDs in African contexts, such as NeuroDev in Kenya and South Africa, expanding knowledge about phenotypic, sociomedical and genetic characteristics relevant in these settings. While there have been huge efforts to bridge the genomic data gap in Africa, challenges still persist with a lack of complete reference genomes and inadequate funding. We hope that this body of work demonstrates the global value and continues to motivate the inclusion of underrepresented populations in clinical and genetic studies of NDDs.

## Methods

### NeuroDev Procedures

Primary behavioral diagnoses in NeuroDev (e.g. autism; attention deficit hyperactivity disorder (ADHD)) were based on DSM-5 criteria and trained clinician judgement. We supplement those diagnoses across all children using several behavioral measures: the Child Behavior Checklist^16^ for ages 1½-5 and 6-18 years (CBCL1/½-5 and CBCL 6-18); the Swanson, Nolan, and Pelham Rating Scale^17^ (SNAP-IV) to measure ADHD symptomatology; the short version of the Developmental, Dimensional and Diagnostic Interview^18,19^ (3di-sv) and the Social Communication Disorders Checklist^20^ (SCDC) to measure characteristics of autism and other behavioral domains. As described in Kipkemoi et al. (2023), we modified several of those measures to better match NeuroDev families’ spoken languages and lived environments.

### Phenotype analyses

All NeuroDev phenotypes were annotated with Human Phenotype Ontology (HPO) terms, a structured ontology which aids in integrating clinical data and conducting clinical genetic analysis. We extracted HPO terms from the Neuromedical Assessment (NMA), CBCL1/½-5 and CBCL 6-18), Modified Checklist for Autism in Toddlers, Revised (MCHAT-R) and Revised with Follow-up (MCHAT-RF)^32^, MAS, and SCDC. For NeuroDev probands from South Africa, HPO terms were also extracted from medical record data. In total, we systematically extracted 235 distinct HPO terms, each of which was mapped back to ancestral terms using the R package “OntologyX”.^33^

We first carried out unadjusted univariable logistic regression on prenatal and perinatal risk factors, comparing children with NDDs and neurotypical children. To evaluate which factors were independently associated with NDDs after accounting for the potential intercorrelations between factors, we carried out a multivariable logistic regression analysis. Risk factors with an adjusted p-value reaching 0.25 in the univariate analysis were included in the multivariable analysis. The lenient significance level used here was to capture as many potentially important factors (clinically and statistically significant) as possible to include in the multivariable analysis. The adjusted models control for parental education, and child age and sex. Sociodemographic factors such as parental education have been strongly linked to child health and development outcomes globally^34^, more specifically higher parental education is associated with an earlier autism diagnosis, a possible reflection of better access to autism services.^35–37^ To address the issue of multiple comparisons, we used the Benjamini-Hochberg Procedure.^38^ After performing the necessary statistical tests to assess the association between each potential risk factor and the outcome of interest in the unadjusted univariable analysis, we obtained a set of p-values representing the significance of these associations. Specifically, we sorted the obtained p-values in ascending order and calculated the critical value corresponding to our desired FDR level (typically set at 0.05). Then, we compared each p-value to its corresponding critical value and considered it significant if it fell below this threshold. Analyses were conducted in R statistical software (version 4.2.2) (R Development Core Team, 2022: https://www.r-project.org/) and STATA version 15.0 (StataCorp LP, College Station, Texas, United States of America [USA]).

### Genetic analyses

#### Exome sequencing and processing

Exome sequencing and data processing were performed by the Genomics Platform at the Broad Institute of MIT and Harvard, following the methods previously described in Kipkemoi et al., 2023. In brief, single nucleotide variants (SNVs) and insertions/deletions (indels) were called using Genome Analysis Toolkit (GATK) HaplotypeCaller package version 3.5. Copy number variants (CNVs) were called from the exome sequencing data following GATK-gCNV best practices. Both the SNV/indel and CNV callsets were uploaded to the *seqr* ^23^ platform on AnVIL for further annotation and analysis by the Broad Institute’s Center for Mendelian Genomics (CMG). The exome sequencing data for each NeuroDev family was analyzed following the protocol previously outlined in Kipkemoi et al., 2023, and all potential causal variants were evaluated following standards established by the American College of Medical Genetics and Genomics/Association for Molecular Pathology (ACMG/AMP).^39^

#### Principal component analysis (PCA)

We utilized the GWASpy pipeline’s (https://github.com/atgu/GWASpy) pre-imputation QC module for variant-level quality control (QC), employing default filters such as minor allele frequency (MAF), call rate, Mendelian error rate, sex checks, inbreeding coefficient, and Hardy-Weinberg disequilibrium. To visualize genetic diversity within the NeuroDev samples, we conducted a principal component analysis (PCA) on array data using the PCA module in GWASpy. This module applied LD-pruning, relatedness estimates, and other necessary filters. For population PCA plots, we used the joint Human Genome Diversity Project and 1000 Genomes Project reference as a global reference, and the African Genome Variation Project as the subcontinental reference. We performed projection PCA, where the reference data defines the principal components, and NeuroDev data is projected onto that space, to mitigate relatedness effects from our family-based data.

#### Rare variant count analyses

Loss-of-function (LoF), missense, in-frame indel, and synonymous variants were identified by annotation with the Ensembl Variant Effect Predictor (VEP) [PMID:27268795] and extracted from the exome sequencing data variant call format (VCF) file using the MANE Select transcript. Only variants with genotype quality ≥40, depth ≥10, allele balance ≥0.2, and without GATK Variant Quality Score Recalibration (VQSR) flags [PMID:20644199] were retained. Variants were annotated with the total allele frequency (AF) and “GroupMax Filtering Allele Frequency” (Grpmax) from the gnomAD reference population database (v4.1) [PMID: 38057664] and only rare variants with an AF <0.001 were retained. The number of rare variants were calculated across all protein-coding genes. These analyses included data from the NeuroDev study, as well as data from other rare disease families sequenced by the Broad Center for Mendelian Genomics (CMG). Genetic ancestry was imputed using the principal component analysis and random forest model used for the Genome Aggregation Database (gnomAD).^40^ The number of non-NeuroDev project probands per ancestry was capped at 50, and only ancestry groups with ≥5 probands were included in the analysis.

#### Family-based exome sequencing analysis

The exome sequencing data for each NeuroDev family was analyzed in *seqr*, following the protocol previously outlined in Kipkemoi et al, 2023. In short, the standard de novo/dominant and recessive variant searches were deployed and potential causal variants were evaluated following standards established by American College of Medical Genetics and Genomics (ACMG)/Association for Molecular Pathology (AMP)/Clinical Genome Resource (ClinGen). For variants impacting genes associated with well-established human conditions, NeuroDev phenotype data was compared to the previously reported clinical presentations of the conditions, bearing in mind potential deviations from expectation due to ancestry. Candidate variants identified in genes of uncertain significance were submitted to the Matchmaker Exchange (MME) network through *seqr* to gather additional support for disease causality.

#### Ethical approval

Written consent for participation was secured from the parents or legal guardians of all children enrolled in NeuroDev, and additional consent was sought from parents for their own participation. Assent was also sought from children above 13 years who were cognitively able to sign a developmentally appropriate assent form. Approval was granted by the Scientific Ethics and Review Unit (KEMRI/SERU/CGMR-C/104/3629) in the KEMRI-Wellcome Trust Research Programme, Kilifi, Kenya, and by the Health Research Ethics Committee (HREC REF:810/2016) in the University of Cape Town, South Africa. In Boston, United States, ethical approval was received from the Harvard T.H. Chan School of Public Health (IRB17-1260 and IRB17-0600).

## Data and code availability

Data and/or research tools used in the preparation of this manuscript were obtained from the National Institute of Mental Health (NIMH) Data Archive (NDA). NDA is a collaborative informatics system created by the National Institutes of Health to provide a national resource to support and accelerate research in mental health. Dataset identifier(s): https://dx.doi.org/10.15154/616g-vf61. This manuscript reflects the views of the authors and may not reflect the opinions or views of the NIH or of the Submitters submitting original data to NDA. All reference panels used in the paper are public resources. Candidate genes identified in the NeuroDev cohort by the Broad CMG have been submitted to Matchmaker Exchange (https://www.matchmakerexchange.org/). Variant classifications have been submitted to ClinVar under submission IDs SUB16040776 and SUB16040807.

## Supporting information

NeuroDev Supplemental Figure 1

NeuroDev Supplemental Table 1

## Data Availability

Data and/or research tools used in the preparation of this manuscript were obtained from the National Institute of Mental Health (NIMH) Data Archive (NDA). NDA is a collaborative informatics system created by the National Institutes of Health to provide a national resource to support and accelerate research in mental health.

## Acknowledgements

We are extremely grateful to the family members for participating in this research. We are grateful to past and present NeuroDev team members, the Neuroethics team members, the clinical laboratories and biobank teams at KEMRI-Wellcome Trust and the University of Cape Town, the community liaison group and neuro-epilepsy clinic at the KEMRI-Wellcome Trust, and developmental and allied clinics at Red Cross Memorial Hospital. We acknowledge James Swanson, Edith Nolan, William Pelham, and team for the use of the SNAP-IV; David Skuse, Richard Warrington, Will Mandy, and team for the use of the 3Di and SCDC; the ASEBA team for the use of the CBCL; Christopher Molteno and team for the use of the Molteno Adapted Scales; and the John C. Raven, John H. Court, and team for the use of the Raven’s Progressive Matrices; Diana Robbins, Deborah Fein, Marianne Barton and team for the use of the MCHAT-R/F.

NeuroDev is supported by the Stanley Center for Psychiatric Research at the Broad Institute. NeuroDev was also supported by a grant from the Simons Foundation or the Simons Foundation International (599648, E.B.R). Research reported in this publication was supported by the: National Institute Of Mental Health of the National Institutes of Health under Award Number U01MH119689; Eunice Kennedy Shriver National Institute Of Child Health & Human Development of the National Institutes of Health under Award Number R01HD102975; National Human Genome Research Institute of the National Institutes of Health under Award Number R01HG012781. The content is solely the responsibility of the authors and does not necessarily represent the official views of the National Institutes of Health. Sequencing was provided by the Broad Institute of MIT and Harvard Center for Mendelian Genomics (Broad CMG) and was funded by the National Human Genome Research Institute; the National Eye Institute; the National Heart, Lung, and Blood Institute grant UM1HG008900; and in part by National Human Genome Research Institute grant R01HG009141.

## Author contributions

E.B.R., K.A.D., A.A., and C.N. conceptualized and designed the study. Z.B., S.M., F.K., C.F., J.R., M.Z., B.C., M.K., A.N., B.Mk., R.M., J.N., S.M., were involved in data collection. B.Me., P.M., C.K., S.B., W.B. were involved in the curation of the data, E.O, M.A., S.L.S, P.K., H.A.K, I.O-O., K.A.R., G.E.V, S.H., K.E.S, J.F., were involved in the analysis of the data. P.K., E.O., S.L.S and E.B.R. wrote the manuscript with input from all authors. P.K., E.E., A.G., K.M. and E.E. were involved in project administration, and C.vdM., C.A.-T., H.B., A.L., D.G.M., A.S.-J., M.S.-B., M.E.T., C.N., A.D.-L., A.A., K.A.D., and E.B.R. supervised various aspects of the project and the core project teams.

## Supplemental information

Supplementary Figure 1 Comparison of rare variant counts in cases and controls by maternal grandmother spoken language Supplementary Table 1 NeuroDev Cohort variant findings

## References

1. Bakare, M.O., Munir, K.M., and Bello-Mojeed, M.A. (2014). Public health and research funding for childhood neurodevelopmental disorders in Sub-Saharan Africa: a time to balance priorities. Healthc Low Resour Settings 2, 2014.1559. 10.4081/hls.2014.1559.

2. Levels and Trends in Child Mortality Report 2018 | UNICEF (2018). https://www.unicef.org/reports/levels-and-trends-child-mortality-report-2018.

3. de Menil, V., Hoogenhout, M., Kipkemoi, P., Kamuya, D., Eastman, E., Galvin, A., Mwangasha, K., de Vries, J., Kariuki, S.M., Murugasen, S., et al. (2019). The NeuroDev Study: Phenotypic and Genetic Characterization of Neurodevelopmental Disorders in Kenya and South Africa. Neuron 101, 15–19. 10.1016/j.neuron.2018.12.016.

4. Kipkemoi, P., Kim, H.A., Christ, B., O’Heir, E., Allen, J., Austin-Tse, C., Baxter, S., Brand, H., Bryant, S., Buser, N., et al. (2023). Phenotype and genetic analysis of data collected within the first year of NeuroDev. Neuron 111, 2800–2810.e5. 10.1016/j.neuron.2023.06.010.

5. Echeverry-Quiceno, L.M., Candelo, E., Gómez, E., Solís, P., Ramírez, D., Ortiz, D., González, A., Sevillano, X., Cuéllar, J.C., Pachajoa, H., et al. (2023). Population-specific facial traits and diagnosis accuracy of genetic and rare diseases in an admixed Colombian population. Sci Rep 13, 6869. 10.1038/s41598-023-33374-x.

6. Africa, S.S. (2025). Education | Statistics South Africa. https://www.statssa.gov.za/?cat=16.

7. Jeste, D.V., Palmer, B.W., Appelbaum, P.S., Golshan, S., Glorioso, D., Dunn, L.B., Kim, K., Meeks, T., and Kraemer, H.C. (2007). A new brief instrument for assessing decisional capacity for clinical research. Arch Gen Psychiatry 64, 966–974. 10.1001/archpsyc.64.8.966.

8. Kipkemoi, P., Mufford, M.S., Akena, D., Alemayehu, M., Atwoli, L., Chibnik, L.B., Gelaye, B., Gichuru, S., Kariuki, S.M., Koenen, K.C., et al. (2024). Evaluation of the psychometric properties of the UBACC questionnaire in a multi-country psychiatric study in Africa. Comprehensive Psychiatry 135, 152526. 10.1016/j.comppsych.2024.152526.

9. Kuja-Halkola, R., Larsson, H., Lundström, S., Sandin, S., Chizarifard, A., Bölte, S., Lichtenstein, P., and Frans, E. (2019). Reproductive stoppage in autism spectrum disorder in a population of 2.5 million individuals. Molecular Autism 10, 45. 10.1186/s13229-019-0300-6.

10. Raven, J. (2000). The Raven’s progressive matrices: change and stability over culture and time. Cogn Psychol 41, 1–48. 10.1006/cogp.1999.0735.

11. Molteno, C.D., Hollingshead, J., Moodie, A.D., Bradshaw, D., Bowie, M.D., and Willoughby, W. (1991). Preschool development of coloured children in Cape Town. South African medical journal = Suid-Afrikaanse tydskrif vir geneeskunde 79, 665–670.

12. Kitsao-Wekulo, P.K., Holding, P.A., Taylor, H.G., Abubakar, A., and Connolly, K. (2013). Neuropsychological Testing in a Rural African School-Age Population: Evaluating Contributions to Variability in Test Performance. Assessment 20, 776–784. 10.1177/1073191112457408.

13. van der Merwe, I., de Klerk, W., and Erasmus, P. (2022). Intelligence Instruments Applied to South African School Learners: A Critical Review. Front. Psychol. 13. 10.3389/fpsyg.2022.853239.

14. Springer, P.E., Laughton, B., Esterhuizen, T.M., Slogrove, A.L., and Kruger, M. (2022). The Molteno Adapted Scale: A child development screening tool for healthcare settings. African Journal of Psychological Assessment 4, 7. 10.4102/ajopa.v4i0.92.

15. Honeth, I., Laughton, B., Springer, P.E., Cotton, M.F., and Pretorius, C. (2019). Diagnostic accuracy of the Molteno Adapted Scale for developmental delay in South African toddlers. Paediatrics and International Child Health 39, 132–138. 10.1080/20469047.2018.1528754.

16. Achenbach, T.M., Dumenci, L., and Rescorla, L.A. (2001). Ratings of relations between DSM-IV diagnostic categories and items of the CBCL/6-18, TRF, and YSR. Burlington, VT: University of Vermont, 1–9.

17. Swanson, J.M., Schuck, S., Porter, M.M., Carlson, C., Hartman, C.A., Sergeant, J.A., Clevenger, W., Wasdell, M., McCleary, R., Lakes, K., et al. (2012). Categorical and Dimensional Definitions and Evaluations of Symptoms of ADHD: History of the SNAP and the SWAN Rating Scales. Int J Educ Psychol Assess 10, 51–70.

18. Skuse, D., Warrington, R., Bishop, D., Chowdhury, U., Lau, J., Mandy, W., and Place, M. (2004). The Developmental, Dimensional and Diagnostic Interview (3di): A Novel Computerized Assessment for Autism Spectrum Disorders. Journal of the American Academy of Child & Adolescent Psychiatry 43, 548–558. 10.1097/00004583-200405000-00008.

19. Kipkemoi, P., Kariuki, S.M., Gona, J., Mwangi, F.W., Kombe, M., Kipkoech, C., Murimi, P., Mandy, W., Warrington, R., Skuse, D., et al. (2024). Utility of the 3Di short version in the identification and diagnosis of autism in children at the Kenyan coast. Front. Psychiatry 15. 10.3389/fpsyt.2024.1234929.

20. Skuse, D.H., Mandy, W.P.L., and Scourfield, J. (2005). Measuring autistic traits: heritability, reliability and validity of the Social and Communication Disorders Checklist. The British Journal of Psychiatry 187, 568–572. 10.1192/bjp.187.6.568.

21. Gurdasani, D., Carstensen, T., Tekola-Ayele, F., Pagani, L., Tachmazidou, I., Hatzikotoulas, K., Karthikeyan, S., Iles, L., Pollard, M.O., Choudhury, A., et al. (2015). The African Genome Variation Project shapes medical genetics in Africa. Nature 517, 327–332. 10.1038/nature13997.

22. Schmitz, M.J., Bashar, A., Soman, V., Nkrumah, E.A.F., Al Mulla, H., Darabi, H., Wang, J., Kiehl, P., Sethi, R., Dungan, J., et al. (2025). Leveraging diverse genomic data to guide equitable carrier screening: Insights from gnomAD v.4.1.0. The American Journal of Human Genetics 112, 181–195. 10.1016/j.ajhg.2024.11.004.

23. Pais, L.S., Snow, H., Weisburd, B., Zhang, S., Baxter, S.M., DiTroia, S., O’Heir, E., England, E., Chao, K.R., Lemire, G., et al. (2022). seqr: A web-based analysis and collaboration tool for rare disease genomics. Hum Mutat. 10.1002/humu.24366.

24. Gona, J., Newton, C., Rimba, K., Mapenzi, R., Kihara, M., Vijver, F., and Abubakar, A. (2016). Challenges and coping strategies of parents of children with autism on the Kenyan coast. Rural Remote Health 16, 3517.

25. Philippakis, A.A., Azzariti, D.R., Beltran, S., Brookes, A.J., Brownstein, C.A., Brudno, M., Brunner, H.G., Buske, O.J., Carey, K., and Doll, C. (2015). The Matchmaker Exchange: a platform for rare disease gene discovery. Human mutation 36, 915–921.

26. Gurovich, Y., Hanani, Y., Bar, O., Nadav, G., Fleischer, N., Gelbman, D., Basel-Salmon, L., Krawitz, P.M., Kamphausen, S.B., Zenker, M., et al. (2019). Identifying facial phenotypes of genetic disorders using deep learning. Nat Med 25, 60–64. 10.1038/s41591-018-0279-0.

27. Bentley, A.R., Callier, S.L., and Rotimi, C.N. (2020). Evaluating the promise of inclusion of African ancestry populations in genomics. npj Genom. Med. 5, 1–9. 10.1038/s41525-019-0111-x.

28. Chen, S., Francioli, L.C., Goodrich, J.K., Collins, R.L., Kanai, M., Wang, Q., Alföldi, J., Watts, N.A., Vittal, C., Gauthier, L.D., et al. (2024). A genomic mutational constraint map using variation in 76,156 human genomes. Nature 625, 92–100. 10.1038/s41586-023-06045-0.

29. Goldenberg, R.L., McClure, E.M., and Saleem, S. (2018). Improving pregnancy outcomes in low- and middle-income countries. Reprod Health 15, 88. 10.1186/s12978-018-0524-5.

30. Keenan, K., Hipwell, A., McAloon, R., Hoffmann, A., Mohanty, A., and Magee, K. (2017). Concordance between maternal recall of birth complications and data from obstetrical records. Early Hum Dev 105, 11–15. 10.1016/j.earlhumdev.2017.01.003.

31. Kesmodel, U.S. (2018). Information bias in epidemiological studies with a special focus on obstetrics and gynecology. Acta Obstetricia et Gynecologica Scandinavica 97, 417–423. 10.1111/aogs.13330.

32. Robins, D.L., Casagrande, K., Barton, M., Chen, C.-M.A., Dumont-Mathieu, T., and Fein, D. (2014). Validation of the Modified Checklist for Autism in Toddlers, Revised With Follow-up (M-CHAT-R/F). Pediatrics 133, 37–45. 10.1542/peds.2013-1813.

33. Greene, D., Richardson, S., and Turro, E. (2017). ontologyX: a suite of R packages for working with ontological data. Bioinformatics 33, 1104–1106. 10.1093/bioinformatics/btw763.

34. Mensch, B.S., Chuang, E.K., Melnikas, A.J., and Psaki, S.R. (2019). Evidence for causal links between education and maternal and child health: systematic review. Tropical Medicine & International Health 24, 504–522. 10.1111/tmi.13218.

35. Dong, H.-Y., Feng, J.-Y., Li, H.-H., Yue, X.-J., and Jia, F.-Y. (2022). Non-parental caregivers, low maternal education, gastrointestinal problems and high blood lead level: predictors related to the severity of autism spectrum disorder in Northeast China. BMC Pediatrics 22, 11. 10.1186/s12887-021-03086-0.

36. Aylward, B.S., Gal-Szabo, D.E., and Taraman, S. (2021). Racial, Ethnic, and Sociodemographic Disparities in Diagnosis of Children with Autism Spectrum Disorder. Journal of Developmental & Behavioral Pediatrics 42, 682. 10.1097/DBP.0000000000000996.

37. Durkin, M.S., Maenner, M.J., Baio, J., Christensen, D., Daniels, J., Fitzgerald, R., Imm, P., Lee, L.-C., Schieve, L.A., Van Naarden Braun, K., et al. (2017). Autism Spectrum Disorder Among US Children (2002–2010): Socioeconomic, Racial, and Ethnic Disparities. Am J Public Health 107, 1818–1826. 10.2105/AJPH.2017.304032.

38. Benjamini, Y., and Hochberg, Y. (1995). Controlling the False Discovery Rate: A Practical and Powerful Approach to Multiple Testing. Journal of the Royal Statistical Society: Series B (Methodological) 57, 289–300. 10.1111/j.2517-6161.1995.tb02031.x.

39. Richards, S., Aziz, N., Bale, S., Bick, D., Das, S., Gastier-Foster, J., Grody, W.W., Hegde, M., Lyon, E., Spector, E., et al. (2015). Standards and guidelines for the interpretation of sequence variants: a joint consensus recommendation of the American College of Medical Genetics and Genomics and the Association for Molecular Pathology. Genetics in Medicine 17, 405–424. 10.1038/gim.2015.30.

40. Karczewski, K.J., Francioli, L.C., Tiao, G., Cummings, B.B., Alföldi, J., Wang, Q., Collins, R.L., Laricchia, K.M., Ganna, A., Birnbaum, D.P., et al. (2020). The mutational constraint spectrum quantified from variation in 141,456 humans. Nature 581, 434–443. 10.1038/s41586-020-2308-7.

