## Supplementary material for "NeuroDev: etiology and experience of neurodevelopmental disorders in Kenya and South Africa": NeuroDev Supplemental Figure 1

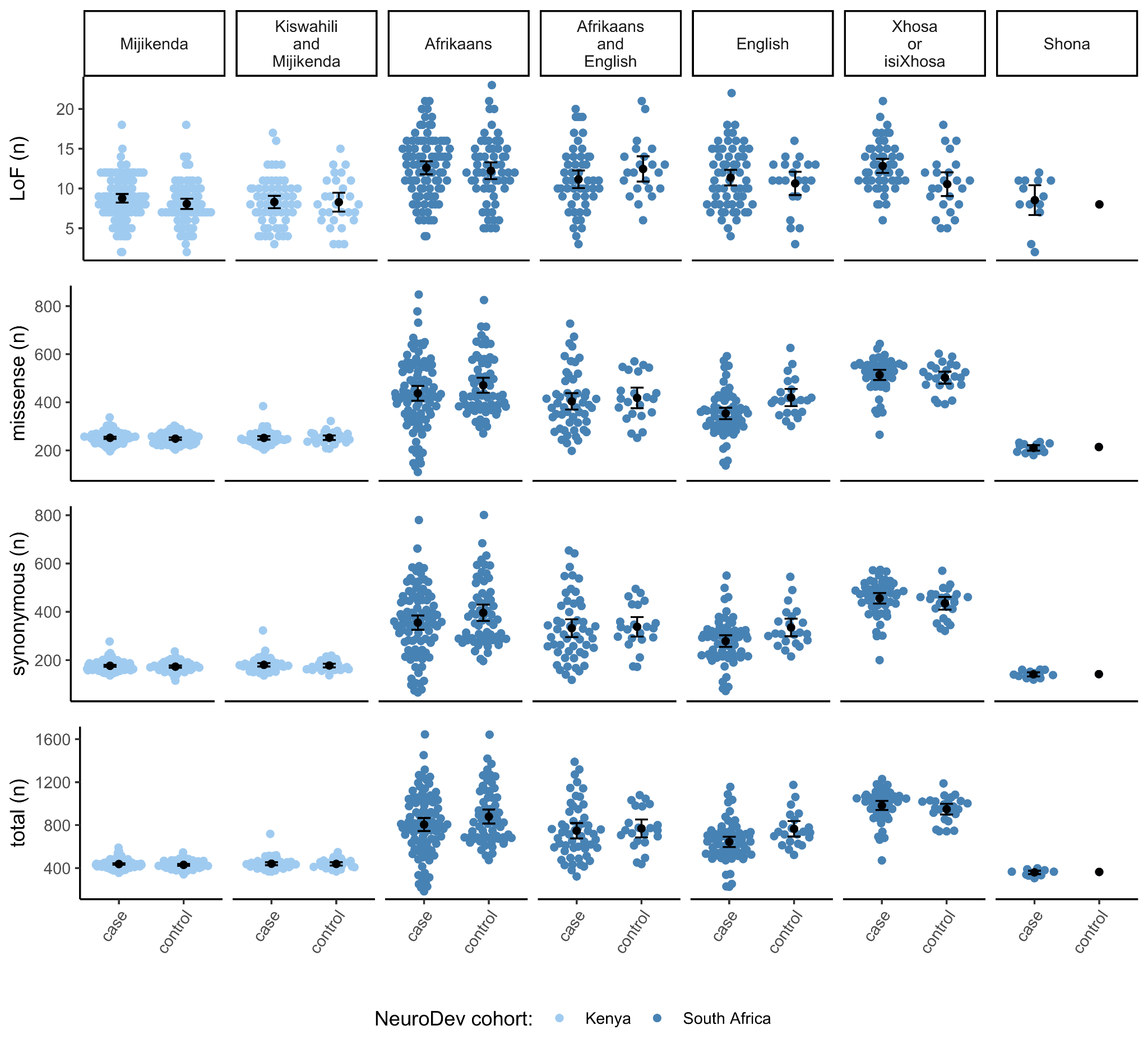

**Supplemental Figure 1. Comparison of rare variant counts in cases and controls by maternal grandmother spoken language.** Rare (allele frequency <0.1% in gnomAD v4.1) variant count of loss-of-function (LoF), missense, and synonymous variants across protein-coding genes between NeuroDev probands and controls by maternal grandmother spoken language. Probands with unknown maternal grandmother spoken language and probands from groups with less than 5 individuals were removed from the analysis. The Shona group only has one control available. This group was kept in the analysis to demonstrate the substantial difference in rare variant rate between this and other maternal grandmother spoken language groups from South Africa that could confound analyses of rare variants rates across phenotypes and environmental exposures.
